# resaCPE: A Rapid, Low-Cost Colourimetric Assay for the Detection of Carbapenemase-Producing Enterobacterales

**DOI:** 10.1101/2025.07.10.25331115

**Authors:** Mitchell A. Jeffs, Gabhan Chalmers, Prameet M. Sheth, Nicole Ricker, Christopher T. Lohans

## Abstract

The use of carbapenem antibiotics is threatened by the global spread of carbapenemase-producing Enterobacterales (CPE), bacterial pathogens which hydrolyze these last-resort antimicrobials. Rapid detection of CPE is vital to ensure timely administration of antimicrobial therapy to infected patients, as well as the implementation of infection control measures to prevent outbreaks in healthcare settings. In this study, we report the development of resaCPE, a rapid, low-cost CPE detection method that couples the inactivation of an imipenem disk with a cell viability assay employing a carbapenem-hyper-susceptible *Escherichia coli* strain. Results are interpreted by a simple colourimetric readout in which CPE-positive samples turn pink, while CPE-negative samples remain purple. The assay was validated with a panel of 91 CPE and non-CPE isolates, and its performance compared to two widely used CPE detection strategies, the modified carbapenem inactivation method (mCIM) and the CARBA-NP test. The resaCPE test demonstrated the same level of performance as the mCIM (100 % sensitivity and specificity), while providing results in significantly less time (3.5 h vs. 20-24 h, respectively). We also observed that the resaCPE test outperformed the CARBA-NP test (95.3 % sensitivity, 100 % specificity), specifically when applied to the detection of CPE isolates producing carbapenemases with weaker hydrolytic activity. Due to the rapid turnaround time, minimal setup requirements, and low cost (∼$1 USD/sample), the resaCPE test is a potentially attractive option for primary CPE screening, particularly in lower resource settings.

## Introduction

Carbapenems (*e.g.,* imipenem, meropenem, ertapenem) belong to the β-lactam class of antibiotics, a group which currently accounts for the majority of antibiotic prescriptions worldwide (1). Carbapenems are active against both Gram-negative and Gram-positive pathogens, and are utilized to treat a broad range of infections, such as pneumonia, urinary tract, bloodstream and intra-abdominal infections (2). These drugs are considered antibiotics of last resort and are held in reserve to target multidrug-resistant bacterial pathogens.

Carbapenem resistance poses an urgent global health threat, endangering the clinical utility of these critical antibiotics. Bacterial pathogens can resist carbapenems through several different mechanisms, including changes to porin levels, modifications to penicillin-binding proteins, efflux and carbapenemase production (3, 4). Carbapenemases belong to the β-lactamase family of enzymes which inactivate β-lactam antibiotics via a hydrolytic mechanism. Carbapenemases are of particular concern, as unlike other types of β-lactamases [*e.g.,* penicillinases, extended-spectrum β-lactamases (ESBLs)], they are capable of hydrolyzing carbapenems. The Ambler classification system divides these enzymes into several groups, namely class A (*e.g.,* KPCs), class B (*e.g.,* NDMs, IMPs, VIMs) and class D (*e.g.,* OXAs) (5). Class A and D enzymes utilize an active site serine residue to catalyze hydrolysis of the β-lactam ring while enzymes in class B employ zinc ions to facilitate hydrolysis. Carbapenemases are produced by many prevalent Gram-negative pathogens; of these, carbapenemase-producing Enterobacterales (CPE; *e.g., Citrobacter freundii, Escherichia coli*, *Klebsiella* spp*., Enterobacter cloacae*) are among the most common and are considered to be a critical threat by the World Health Organization (6).

Treatment options for infections caused by CPE are limited, as many carbapenemases can hydrolyze nearly all clinically available β-lactam antibiotics. Additionally, genes that encode for carbapenemases are often located within mobile genetic elements, which may also carry resistance genes for other classes of antibiotics, and which can be spread via horizontal gene transfer (7–11). Thus, rapid detection of CPE is vital to ensure timely administration of appropriate antibiotic therapy. Additionally, specific infection prevention and control (IPAC) measures are typically implemented for patients who test positive for CPE infection or colonization so as to prevent outbreaks (12, 13). These measures may include patient isolation in private rooms, increased use of personal protective equipment (PPE) by hospital staff, and increased room disinfection. Outbreak prevention is vital, as CPE infections are associated with high mortality rates (14–16). Implementation of surveillance programs (*e.g.,* using diagnostics to detect CPE) ensures that such IPAC practices can be implemented rapidly when patients test positive for CPE, limiting transmission within healthcare settings.

There are several methods currently employed by clinical microbiology labs for the detection of CPE. Molecular detection of carbapenemase-encoding genes by polymerase chain reaction (PCR) has become the gold standard (17). However, despite high sensitivity and specificity, and short turnaround times, implementation of PCR-based assays may be limited in lower resource settings due to the high costs associated with the equipment and reagents used to run these tests (18). Additionally, these methods may not detect novel or rare carbapenemase genes, as PCR panels typically only target the most commonly encountered carbapenemases.

In addition to molecular assays, phenotypic assays are also available for the detection of CPE in clinical settings. The modified carbapenem inactivation method (mCIM) has been endorsed by the Clinical and Laboratory Standards Institute (CLSI), and is used extensively due to its robust performance, low cost and ease-of-use (19–22). However, this assay suffers from long turnaround times as it measures the impact of a meropenem disk on bacterial growth, typically requiring 20 - 24 h to provide a result. Faster phenotypic assays that measure carbapenemase activity have been developed to address the long turnaround times associated with growth-based assays (23). The CARBA-NP test is the sole rapid biochemical assay endorsed by the CLSI, relying on pH changes which result from carbapenemase-catalyzed imipenem hydrolysis (23). Although this test has a drastically reduced turnaround time (∼2 h) compared to the mCIM, some labs have found that the CARBA-NP test exhibits poor sensitivity for carbapenemases with weaker hydrolytic activity (*e.g.,* OXA-48-like) (24–27). This is particularly significant in regions where the prevalence of OXA-48-producers is high, including across much of western Europe, North Africa, parts of the Middle East and India (28). Additionally, the CARBA-NP assay requires labs to prepare pH-adjusted buffer solutions (*i.e.,* CARBA-NP solutions A and B) on a regular basis. The RAPIDEC-CARBA-NP is a commercially available version of the CARBA-NP test that provides all required components, thus increasing ease-of-use (29). However, the RAPIDEC-CARBA-NP is more costly and has also been observed to suffer from the inconsistent detection of OXA-48-like enzymes (26, 27).

Lateral flow assays such as the NG-Test Carba-5 have also been adopted in clinical microbiology labs. This antibody-based assay provides extremely rapid results (15 - 30 min) and demonstrates a high degree of sensitivity and specificity for the “big five” carbapenemases (*i.e.*, OXA-48, KPC, IMP, VIM, NDM) (27, 30, 31). Despite these advantages, the cost per test is high (21), this assay cannot detect all variants of the five carbapenemases (32, 33), and it also cannot detect other types of carbapenemases.

The efficacy of carbapenem antibiotics against CPE has been preserved in part by the development of combination therapies which include β-lactamase inhibitors (*e.g.,* avibactam, vaborbactam). Despite this, carbapenemase variants which are less susceptible to inhibition have begun to emerge (34–37). Additionally, there are currently no inhibitors available for clinical use which can target metallo-β-lactamases (*e.g.,* NDM, IMP, VIM) (38). Although inhibitor efficacy can be evaluated through growth-based assays, current rapid testing methods such as the CARBA-NP or NG-Test Carba-5 tests do not typically incorporate clinically used inhibitors into their workflows. As resistance to β-lactamase inhibitors increases in prevalence, there will be a growing need for rapid CPE detection methods that can also inform on clinical inhibitor effectiveness.

In this study, we developed a colourimetric test which can be applied to the rapid detection of CPE. This assay, the resaCPE test, couples imipenem inactivation by a CPE with a cell viability assay employing resazurin, a dye which changes from purple to pink when it is reduced to resorufin by metabolically active cells. The resaCPE assay was validated with a diverse panel of 91 CPE and non-CPE isolates, yielding a sensitivity and specificity of 100 %. The resaCPE test exhibited the same sensitivity and specificity as the mCIM while providing same-day results and outperformed the CARBA-NP test (95.3 % sensitivity, 100 % specificity) when applied to the detection of CPE isolates which produce carbapenemases with weaker hydrolytic activity (*e.g.,* OXA-48-like). Additionally, carbapenemase inhibitors such as avibactam could be easily incorporated into the assay workflow to evaluate inhibitor efficacy against a particular CPE. The resaCPE test requires minimal setup and does not depend on specialized lab equipment, thus it could be suitable for implementation in clinical microbiology labs. Additionally, this test is low-cost (∼$1 USD/sample) and may be particularly attractive for lower resource settings.

## Materials and Methods

### Reagents

BD BBL cation-adjusted Mueller-Hinton broth (CAMHB), trypticase soy broth (TSB), proteinase K, Dulbecco’s phosphate-buffered saline (DPBS), resazurin sodium salt, zinc sulfate and phenol red were purchased from Fisher Scientific. Bacterial protein extraction reagent B-PER was purchased from Thermo Fisher. Imipenem (10 µg) and meropenem (10 µg) disks were purchased from Hardy Diagnostics. Avibactam was purchased from Medkoo Biosciences.

### Bacterial Strains

*E. coli* BW25113 (National BioResource Project, NBRP) was transformed with pACYC184 vectors carrying genes encoding for an ESBL (CTX-M-15) and carbapenemases (NDM-1, IMP-1, KPC-2, OXA-48) as previously reported (39). The native promoter and gene encoding OXA-10 was amplified from the genetic material of *Klebsiella pneumoniae* AR0041 by PCR and cloned into the HindIII site of pACYC184 using the NEBuilder HiFi DNA Assembly kit (New England Biolabs). The sequence of the recombinant plasmid was confirmed by whole plasmid sequencing (Plasmidsaurus).

CPE clinical isolates were collected by the clinical microbiology lab at Kingston Health Sciences Center (KHSC; Kingston, Canada) during the normal course of patient care, and ESBL-producing strains were previously isolated from dairy cattle manure (40, 41). The panel of isolates comprised 65 CPE and 26 ESBL-producing non-CPE belonging to several Enterobacterales species (Tables S1, S2). The CPE isolates were previously tested by the KHSC microbiology lab for carbapenemase production via the mCIM test, and the presence of carbapenemase genes (IMI, NDM, KPC, OXA-48, VIM) was confirmed by multiplex PCR. The presence of ESBL genes (TEM, CTX-M, CMY families) and absence of carbapenemase genes in the non-CPE isolates was verified by whole genome sequencing (40, 41).

All bacterial strains used in this study were cultured for 16 – 20 h at 37 °C on CAMHB agar plates. Carbapenemase-negative control *E. coli* ATCC 25922 and indicator strain *E. coli* BW25113 Δ*ompA* (NBRP) (42) were grown on agar plates lacking antibiotic. Clinical CPE isolates were cultured on agar plates supplemented with 1 µg/mL imipenem, and ESBL-producing non-CPE isolates were grown on agar plates supplemented with 1 µg/mL cefotaxime.

### resaCPE Assay

Test isolates, the non-carbapenemase-producing control *E. coli* ATCC 25922, and the indicator strain *E. coli* BW25113 Δ*ompA* were cultured as described above.

#### Part A (Tube Set 1)

One 5 mL polystyrene culture tube (Falcon) was prepared for each test sample and for the negative control. To each tube, 200 µL of lysis reagent (B-PER + 100 µM ZnSO_4_) was added, and, using a rigid inoculation loop (Fisher Scientific), 2 x 1 µL loopfuls of negative control or test strain colonies were emulsified in their respective tubes. Each tube was vortexed for 10 s. Using sterile forceps, a 10 µg imipenem disk was added to each lysate, and these tubes were set aside until part B was completed.

#### Part B (Tube Set 2)

A second set of 5 mL culture tubes was prepared, with one tube for the negative control, one for each test strain, and one for the viability control. To each tube, 975 µL of sterile CAMHB was added. In a new 5 mL tube, a suspension of the indicator strain was prepared to approximately 0.5 McFarland Units (MFU; OD_600_ ∼ 0.1) in CAMHB. Then, 25 µL of this suspension was transferred to the negative control, test sample, and viability control tubes. Tube sets from both parts A and B were incubated at 37 °C for 90 min without shaking.

#### Part C

After the 90 min incubation, 20 µL of a 500 µg/mL proteinase K solution (prepared in sterile water) was added to each tube in set 2 (final concentration ∼10 µg/mL). The imipenem disks were transferred from the tubes in set 1 to the corresponding tubes in set 2 using a separate sterile 1 µL inoculation loop for each disk. Note that the viability control does not receive a disk. Lastly, 200 µL of a 0.15 mg/mL resazurin solution (prepared in sterile water) was added to each tube in set 2. These tubes were then incubated at 37 °C for 2 h, and the colour of the solutions was visually inspected. A colour change from purple to pink was interpreted as a CPE-positive result, while a lack of colour change was interpreted as a CPE-negative result.

### CARBA-NP Assay

The CARBA-NP test was performed as outlined in the CLSI M100 Edition 34 document (43). CARBA-NP solution A was prepared by adding 2 mL of 0.5 % phenol red and 180 µL of 10 mM ZnSO_4_ to 16.6 mL of sterile water, which was then adjusted to pH 7.8 and stored at 4 °C for up to two weeks. Bacterial strains were cultured on agar plates as described above. CARBA-NP solution B was prepared on the day of the assay by dissolving imipenem in CARBA-NP solution A to a concentration of 3 mg/mL. A 1 µL loopful of each test strain was added to two separate 1.5 mL tubes containing 100 µL of B-PER and vortexed for 5 - 10 s. To the first set of tubes, 100 µL of solution A was added, and to the second set of tubes, 100 µL of solution B was added. These samples were incubated at 37 °C for 2 h without shaking, and colour change was monitored after this incubation period.

### mCIM

The mCIM was performed as outlined in the CLSI M100 Edition 34 document Bacterial strains were cultured on agar plates as described above. On the day of the experiment, a 1 µL loopful of each strain was suspended in 2 mL sterile TSB and vortexed for 5 - 10 s. A 10 µg meropenem disk was added to each tube, which were then incubated at 37 °C for 4 h without shaking. Just prior to the end of the incubation period, a suspension of *E. coli* ATCC 25922 was prepared in sterile TSB to 0.5 MFU. A sterile cotton swab (Puritan) was dipped into the suspension and used to coat the surfaces of CAMHB agar plates lacking antibiotic. The meropenem disks were transferred from the bacterial suspensions to the surfaces of the agar plates using sterile inoculation loops. Plates were inverted, then incubated at 37 °C for 20 h. Results were interpreted as per CLSI guidelines (43).

## Results and Discussion

The resaCPE assay is a colourimetric test which combines an imipenem inactivation step with a cell viability test employing resazurin (Figure 1). In short, colonies of test isolates are first lysed, then incubated with an imipenem disk. This disk is then transferred to a suspension of an imipenem-susceptible indicator strain, to which proteinase K and resazurin are then added. If the test isolate is a CPE, the imipenem is hydrolyzed, allowing the indicator strain to survive this treatment. Consequently, the living indicator strain converts resazurin into a pink product.

**Figure 1:**
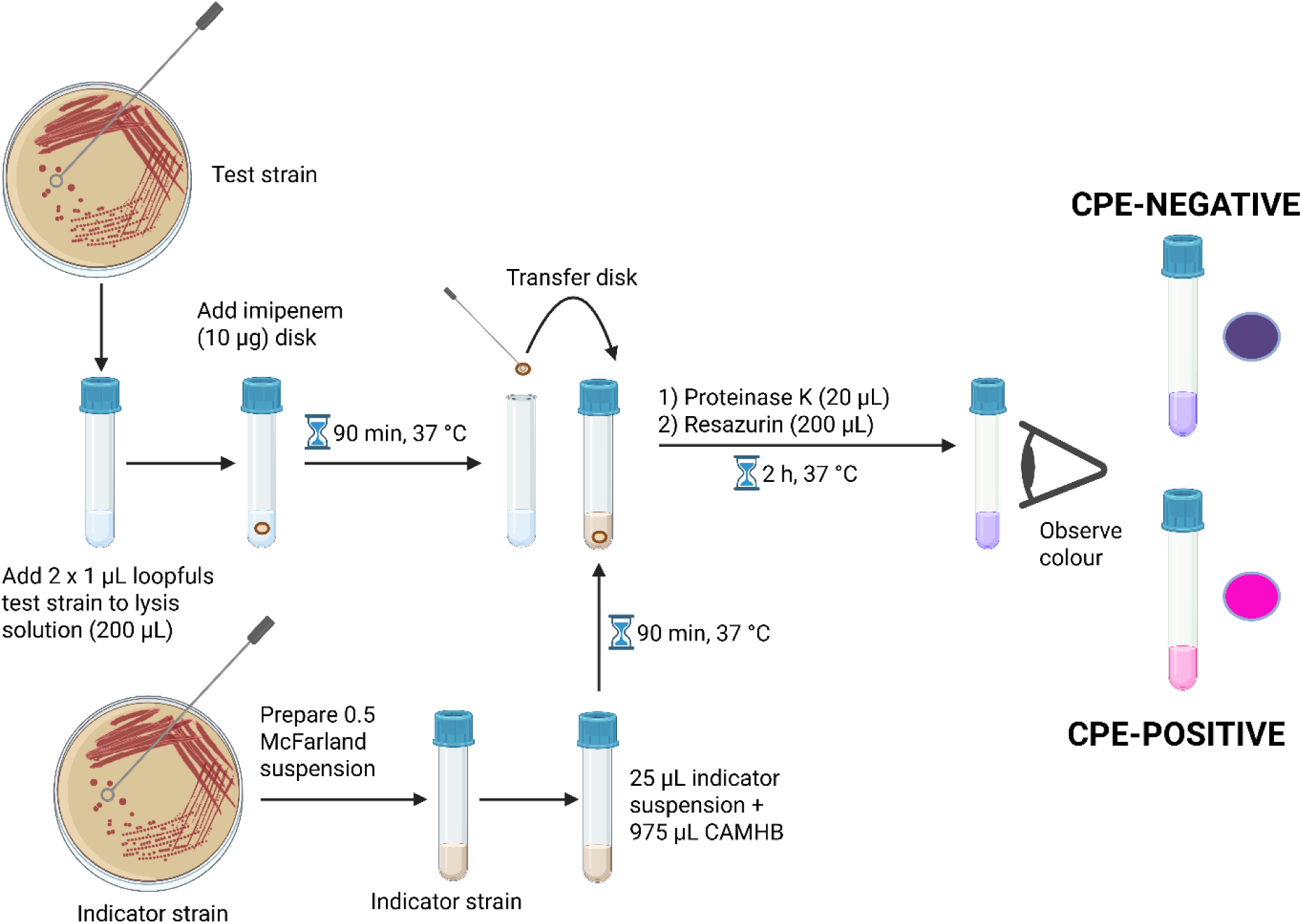
Graphical overview of the resaCPE assay workflow. If the test strain is a CPE, the imipenem is hydrolyzed, and the indicator strain remains viable, resulting in a colour change from purple to pink. If the test strain is not a CPE, the imipenem is not degraded and kills the indicator strain, and no colour change occurs. Created in BioRender. Jeffs, M. (2025) https://BioRender.com/l37bl3z.

It is vital that the indicator strain is susceptible to imipenem, thus ensuring that it does not remain viable following exposure to this antibiotic. If the indicator strain were to survive the imipenem treatment, it would cause an undesirable background colour change which could yield a false positive result even if the test strain is not a CPE. Preliminary resazurin assays were used to compare the susceptibilities of potential indicator strains, including wild-type *E. coli* BW25113 and a series of *E. coli* mutants lacking genes which encode for proteins that stabilize the outer membrane (Δ*ompA,* Δ*lpp,* Δ*nlpI*), to imipenem. In these assays, the *E. coli* Δ*ompA* strain demonstrated the greatest susceptibility to imipenem, and the disruption of *ompA* did not appear to impact the metabolism of resazurin in the absence of imipenem (Figure S1). OmpA is a β-barrel protein which helps to anchor the outer membrane of *E. coli* to the peptidoglycan layer (43). When this protein is absent, the outer membrane is destabilized, increasing bacterial susceptibility to some β-lactam antibiotics (45). Based on these results, the Δ*ompA* mutant was chosen as the indicator strain for the resaCPE assay to minimize potential background colour change during the resazurin reaction resulting from surviving indicator strain cells.

In early experiments employing the resaCPE assay to detect carbapenemases in clinical isolates, we observed that lysates of some test strains had a negative impact on the viability of the indicator strain, decreasing resazurin turnover and colour change (Figure S2A). However, addition of proteinase K to the assay mixture protected the indicator strain from the components of the test strain lysate (Figure S2B). Based on these results, we hypothesized that some of the CPE isolates may produce antimicrobial peptides and proteins such as bacteriocins. These proteins typically target bacterial species closely related to the strain which produces them, although several reports have described the production of broad-spectrum bacteriocins by bacterial pathogens from human and animal sources (46–49). Bacteriocins or other antimicrobial proteins that are produced by a test isolate could kill the indicator strain in the resaCPE assay, thus preventing a colour change and yielding a false negative result. Hence, the addition of proteinase K to the assay appears to overcome this limitation by degrading potential bacteriocins and protecting the indicator strain.

Further optimization with regards to the lysis conditions (Figure S3), the length of time that lysates were incubated with an imipenem disk (Figure S4A), the length of time the indicator strain suspension was incubated prior to the addition of resazurin (Figure S4A), and the volume of indicator suspension used in the resazurin reaction (Figure S4B) was conducted. Following this optimization, the final protocol for the resaCPE assay was validated with a panel of laboratory *E. coli* strains transformed with plasmids encoding for carbapenemases and ESBLs (Figure 2). All carbapenemase-producing strains tested positive, as indicated by a colour change from purple to pink, while the ESBL-producing strains tested negative and did not change colour due to their inability to hydrolyze imipenem.

**Figure 2:**
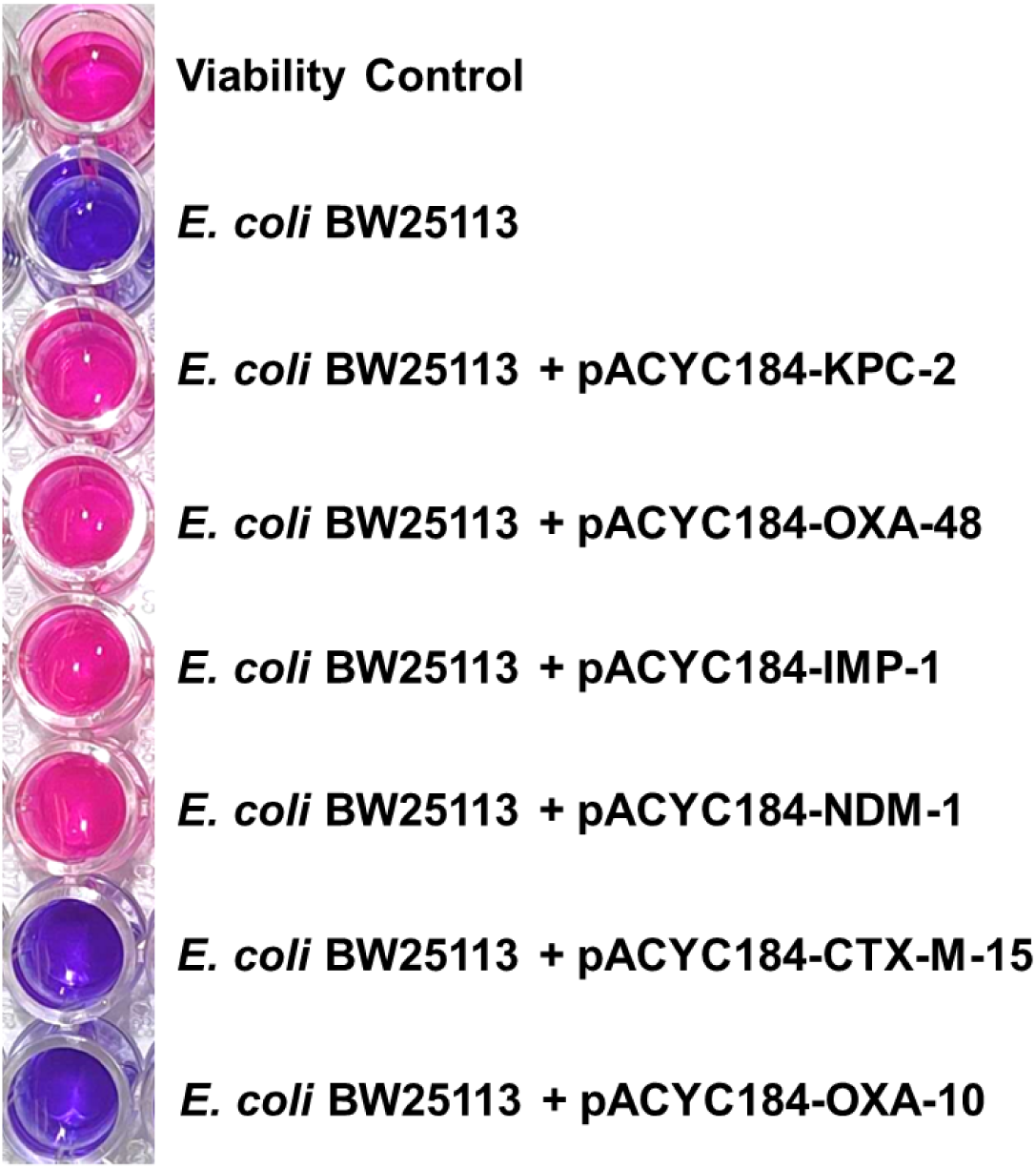
Validation of the resaCPE assay with *E. coli* strains transformed with pACYC184 plasmids carrying genes encoding for carbapenemases and ESBLs. All strains producing carbapenemases (KPC-2, OXA-48, IMP-1, NDM-1) caused a colour change from purple to pink and thus were interpreted as positive results. The two non-carbapenemase-producing strains (*i.e.*, those producing CTX-M-15 or OXA-10) did not cause a colour change and thus yielded negative results. The viability control is used to monitor the progression of colour change driven by the indicator strain in the absence of an imipenem disk.

Next, we challenged the sensitivity of the resaCPE test with a panel of 65 clinical carbapenemase-producing strains, including *C. freundii, E. cloacae*, *E. coli*, *Klebsiella oxytoca*, *Klebsiella pneumoniae* and *Morganella morganii*. Collectively, these isolates produce a diverse range of carbapenemases which are commonly encountered by a clinical microbiology lab (*i.e.*, KPC, IMI, NDM, VIM and OXA-48-like) (Tables 1, S1). All CPE isolates were successfully detected with the resaCPE assay, thus demonstrating a sensitivity of 100 % when challenged with this panel. The performance of the resaCPE assay was compared to that of the CARBA-NP and mCIM tests, which yielded sensitivities of 95.3 % (62/65 positive) and 100 % (65/65 positive), respectively, against the same panel (Tables 1, S1). Notably, the CARBA-NP test failed to detect 2/8 *E. coli* strains producing OXA-48-like carbapenemases, consistent with other reports where poor sensitivity for these carbapenemases was observed (24, 25). In addition, an IMI-producing *E. cloacae* isolate tested false-negative with the CARBA-NP test. All three isolates which yielded false-negative results with the CARBA-NP test exhibited slow hydrolytic rates of imipenem in spectrophotometric analyses (Figure S5).

**Table 1:** Summary of resaCPE, CARBA-NP and mCIM test results for the panel of CPE isolates.

| Ambler Class (n) | Carbapenemase Type | Positive Results |  |  |
| --- | --- | --- | --- | --- |
|  |  | *resaCPE* | CARBA-NP | mCIM |
| Class A (28) | KPC (26) | 26 / 26 | 26 / 26 | 26 / 26 |
|  | IMI (2) | 2 / 2 | 1 / 2 | 2 / 2 |
| Class B (28) | VIM (15) | 15 / 15 | 15 / 15 | 15 / 15 |
|  | NDM (13) | 13 / 13 | 13 / 13 | 13 / 13 |
| Class A + B (1) | KPC + NDM (1) | 1 / 1 | 1 / 1 | 1 / 1 |
| Class D (8) | OXA-48-like (8) | 8 / 8 | 6 / 8 | 8 / 8 |
| **Total** |  | **65 / 65** | **62 / 65** | **65 / 65** |
| **Sensitivity** |  | **100 %** | **95.3 %** | **100 %** |

A panel of 26 ESBL-producing *E. coli* and *Proteus mirabilis* isolates was used to challenge the specificity of the resaCPE test. None of the tested isolates produced a colour change, and so the assay demonstrated a specificity of 100 % against this panel with no false positives (Tables 2, S2). The CARBA-NP and mCIM assays yielded the same results when applied to these isolates (Tables 2, S2).

**Table 2:** Summary of resaCPE, CARBA-NP and mCIM test results for the panel of non-CPE isolates.

| β-lactamase (n) | False-Positive Results | | |
| --- | --- | --- | --- |
|  | resaCPE | CARBA-NP | mCIM |
| CTX-M (19) | 0 / 19 | 0 / 19 | 0 / 19 |
| CTX-M + TEM (4) | 0 / 4 | 0 / 4 | 0 / 4 |
| CMY (2) | 0 / 2 | 0 / 2 | 0 / 2 |
| CMY + CTX-M (1) | 0 / 1 | 0 / 1 | 0 / 1 |
| **Total** | **0 / 26** | **0 / 26** | **0 / 26** |
| **Specificity** | **100 %** | **100 %** | **100 %** |

Following assay validation, we expanded our testing to see if the carbapenemase inhibitor avibactam could be incorporated into the workflow. Current methods for evaluating the effectiveness of β-lactam+β-lactamase inhibitor combinations include broth microdilution assays and E-tests, which suffer from long turnaround times (18 - 24 h). Incorporation of avibactam (32 µg/mL) into the resaCPE test allowed for the successful differentiation of clinical CPE isolates which produce carbapenemases that are inhibited by avibactam (*e.g.*, certain KPC and OXA-48-like enzymes) from those which produce carbapenemases that are not inhibited (*e.g.*, NDM, VIM) (Figure 3). Notably, within the panel of isolates tested, some strains were found to produce KPC and OXA-48-like enzymes which were less susceptible to inhibition by avibactam. These results were corroborated with UV-Vis spectrophotometric analyses measuring the rate of imipenem hydrolysis in the presence and absence of avibactam (Figure S6).

**Figure 3.**
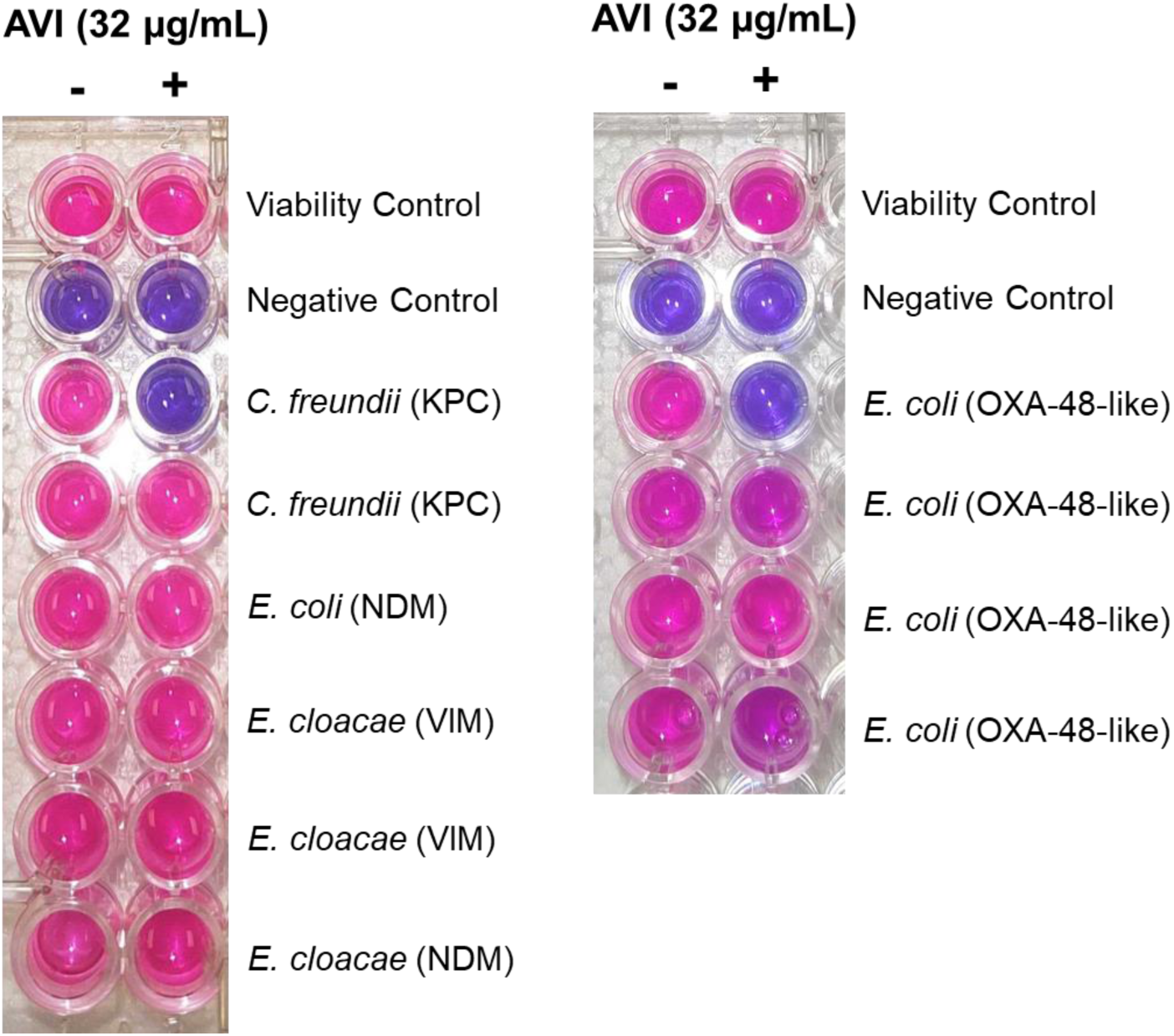
resaCPE assays evaluating avibactam efficacy against a representative selection of CPE isolates. Test strains which did not change colour in the presence of avibactam (AVI) produce carbapenemases which are inhibited by avibactam. Test strains which turn pink in the presence of avibactam produce carbapenemases which are not (or are less) susceptible to avibactam. The viability control is used to monitor the progression of colour change driven by the indicator strain in the absence of an imipenem disk. The negative control strain is *E. coli* ATCC 25922.

Future studies will challenge the resaCPE assay with a larger and more diverse library of isolates, and will evaluate the feasibility of implementing the resaCPE assay in hospital settings. One factor that could impact implementation of the resaCPE test in lower resource settings is the requirement of a -80°C freezer, which is needed to store frozen stocks of the indicator strain. To overcome this, future work will investigate the stability of the indicator strain to lyophilization, overcoming the need for a -80 °C freezer for storage. We will also explore the application of this assay to the direct detection of CPE from rectal swabs. This would negate the need for overnight culturing of potential CPE isolates prior to testing as is typically required for phenotypic CPE testing methods, including the resaCPE test described here.

Herein, we introduce the resaCPE assay, a colourimetric method for the rapid detection of CPE. This proof-of-concept study demonstrates that the resaCPE test can be used to accurately detect the production of carbapenemases by clinical Enterobacterales isolates within 3.5 h and with 100 % sensitivity and specificity. This test is rapid, low cost (∼$1 USD/sample), does not require any specialized equipment and has a simple setup, thus potentially making it suitable for implementation in clinical microbiology labs. Additionally, carbapenemase inhibitors can easily be incorporated into the assay workflow if desired. Rapid detection of CPE will continue to be vital in clinical settings to ensure that patients receive appropriate antimicrobial therapy and so that infection prevention and control decisions can be made efficiently to help prevent the spread of CPE in healthcare settings.

## Data Availability

All data produced in the present work are contained in the manuscript

